# The association between an agricultural intervention integrated within a group-based microfinance program and dietary diversity in rural Kenya

**DOI:** 10.64898/2026.07.23.26358820

**Authors:** Charles Opondo, James Kamadi, James Akiruga Amisi, Ricky Camplain, Sonak Pastakia, Nana Gletsu-Miller, Christina Ludema, Molly Rosenberg

## Abstract

Low dietary diversity is a risk factor for micronutrient deficiencies. Agricultural interventions that support crop production and livestock rearing may promote dietary diversity and achieve better nutritional outcomes, particularly among low-income households. Their effectiveness may be strengthened when integrated within microfinance platforms. The objective of this study was to estimate the association between an agricultural intervention delivered within a group-based microfinance program and dietary diversity in rural Western Kenya, and determined effect modification by sex. We conducted a cross-sectional design from June to August 2025 in two village wards of Webuye sub-County in rural Western Kenya, using a validated dietary diversity scale to assess the total number of food groups consumed in the previous 24 hours. We interviewed 312 microfinance participants, 156 of whom were in agricultural intervention comprised of farm input subsidies and training, while 156 were not in the intervention. We specified modified Poisson models to estimate the association between intervention receipt and dietary diversity, and tested whether the estimated association differed by sex. Participation in the agricultural intervention was associated with higher diversity among microfinance program members [adjusted PR (95% CI): 1.46 (1.02, 2.06)]. We did not find evidence that the association differed by sex (Wald p-value for interaction term=0.83). Participation in an agricultural intervention was associated with improved dietary diversity. Future prospective using large samples studies should build on these findings to establish temporal ordering, and assess the feasibility and sustainability of integrating agricultural support into microfinance platforms to improve dietary diversity in low-resource settings.

## INTRODUCTION

Undernutrition is a major public health concern in sub-Saharan Africa. Undernutrition cases rose from 59 in 2018 to 235 million in 2019 ^(1,2)^. In Kenya, nearly 40% of the Kenyan population have insufficient intake of micronutrients such as vitamin A, zinc, and iron^(3)^. The health consequences of undernutrition are severe and multifaceted, contributing to stunting, anaemia, depression, and an increased mortality risk^(1,4,5)^ . These outcomes not only impact an individual’s well-being but also hinder economic productivity and perpetuate the cycles of poverty.

The causes of undernutrition are interconnected, passed down through generations, and structured hierarchically at proximal and distal levels ^(6)^ . Proximal risk factors of undernutrition include inadequate dietary intake, and infectious diseases. Distal risk factors of undernutrition include household food insecurity, poor infant feeding practices, poor health and sanitation environment; poor housing and living conditions. Even more distal risk factors of undernutrition include gender inequality, lack of access to social protection programs, poor infrastructure, and poverty ^(7)^ . Given the public health importance of undernutrition in low- and middle-income countries, it is critical to evaluate strategies and programs that are designed to address these interconnected risk factors to alleviate poverty, improve food security, and enhance socio-economic conditions in low- and middle-income countries like Kenya.

Agricultural interventions embedded within a group-based microfinance programs may enhance socioeconomic well-being by increasing household income and improving access to diverse food groups among participants. However, the growing body of evidence on the impacts of agricultural interventions on dietary diversity in low- and middle-income countries is inconsistent. Agricultural interventions designed to address undernutrition have improved dietary diversity in Tanzania, the Democratic Republic of Congo, Malawi, Sri Lanka and India ^(8–13)^ . Similarly, programs promoting indigenous vegetable cultivation and livestock rearing have been associated with increased consumption of nutrient dense foods in Nepal, Rwanda, and Myanmar ^(14–16)^ . In contrast, studies from Vietnam and Zambia found no association between agricultural interventions and dietary diversity ^(17,18)^ . The inconsistencies in study findings may stem from variations in program design, methodological approaches, and implementation frameworks, which limit the generalizability of findings to other settings such as Kenya. The inconsistent results and limited transportability of evaluation impacts highlight a persistent knowledge gap in rural Kenyan contexts.

To address this gap, we evaluated the association between an agricultural intervention and dietary diversity in rural western Kenya. The agricultural intervention is embedded in a group-based microfinance, and portable healthcare delivery program; the Bridging Income Generation with grouP Integrated Care (BIGPIC) program, that has been running since 2013 in Western Kenya ^(19,20)^. The agricultural intervention comprises agribusiness training on livestock and poultry rearing,

African indigenous vegetables, as well as provision of discounted prices on agricultural inputs such as fertilizers and seedlings to participants, but it has not been previously evaluated on dietary diversity outcomes. We hypothesized that participants in the BIGPIC microfinance program who received the agricultural intervention would report higher dietary diversity compared to BIGPIC microfinance participants who did not receive the agricultural intervention.

## METHODS

### Study Design and Settings

We conducted a cross-sectional study among households in two neighbouring villages in Milo and Matulo wards in Webuye sub-County of Bungoma County in rural Western Kenya. Each ward had an estimated population of about 15,000 residents at the time of conducting the survey. An estimated 35% of the population in Bungoma County experience poverty, slightly higher than the national average of 32% ^(21)^ . Participants from the two wards had similar sociodemographic characteristics, and earned most of their household income from livestock, dairy, poultry, crop farming and small-scale business activities.

### Study Sample

From a roster of 4545 current members of BIGPIC program, we purposively recruited 156 eligible participants who had received agricultural training and farm input support through the agricultural intervention component of the BIGPIC program. We recruited an additional 156 newly enrolled BIGPIC members, classified as unexposed who had joined the program less than seven months prior to the study period. All participants were aged 18 years or older, residents of Milo or Matulo wards for at least one year prior to the study, and were active members of the BIGPIC microfinance program during the study period. Participants who reported themselves as having received the agricultural intervention but had their exposure information missing from the BIGPIC administrative records were excluded from the study after screening for eligibility.

### Piloting and Data Collection Procedures

Data were collected using tablets programmed with interviewer-administered questionnaires in REDCap ^(22)^ . Prior to data collection, the questionnaire was piloted among five eligible participants to assess face validity, comprehension, and cultural appropriateness, and to allow for necessary revisions. Participants involved in the pilot were excluded from the main study, although they had similar sociodemographic characteristics to the study population.

### BIGPIC Family Program

Milo and Matulo has an established BIGPIC Family program operating within the Academic Model Providing Access to Healthcare (AMPATH) in Western Kenya. The AMPATH is a global consortium of North American Universities, Moi University and Moi Teaching and Referral Hospital, and the Government of Kenya. The BIGPIC Family program provides a platform where microfinance group members meet weekly to take out and repay group-funded loans, and may also access a range of integrated services, including screening, and care for diabetes, hypertension, and mental health. In addition, the program offers group-based primary healthcare delivery during microfinance meetings, business literacy, and socioeconomic strengthening activities such as agricultural interventions. The agricultural component includes training in sustainable farming practices, and provision of farm inputs. These are delivered through training-of-trainers (TOT) model, in which group leaders designated as Group Empowerment Service Providers (GESPs) are trained by county government agricultural extension workers, and subsequently support microfinance group members.

### Agricultural Intervention

We established exposure status using administrative records tracking agricultural training delivered by the GESPs and was further verified through participant self-report about participants’ exposure to the agricultural intervention. The BIGPIC program participants who received the agricultural intervention were classified as exposed (1), while those who did not classified as unexposed (0). The unexposed group consisted of newly enrolled microfinance members who had not received agricultural training or farm input support during the study period. Participants in the exposed group received one cycle of farm input support, and two training cycles covering land preparation, and nursery seedbed preparation for African indigenous vegetables farming, as well as livestock, and poultry rearing. These activities were delivered during each of the two planting seasons in the previous year.

### Dietary Diversity

Our outcome variable was dietary diversity, measured using a validated dietary diversity scale to calculate the number of food groups consumed in the household during the previous 24 hours. The scale uses culturally appropriate foods intended for the general population of the study setting as examples of each food group queried ^(23)^. We assessed participants’ responses to questions: “Yesterday did you and your family eat any of the following food groups? 1.) starch (e.g. Maize ugali, maize porridge, rice, bread, chapati, injera, pasta, or noodles); 2.) fruits (e.g. Pawpaw, mango, passion fruit, or matunda ya damu); 3.) vegetables (e.g. Sukuma wiki, Ethiopian kale, spinach, nightshade, amaranth, saget, or cowpea leaves); 4.) animal proteins (e.g. goat meat, beef, minced beef, mutton, liver or matumbo), and 5.) pulses, nuts or seeds. Participants who reported that their household consumed all five food groups within the recall period of 24 hours were classified as having a high dietary diversity, while those who’s households consumed less than five food groups were classified as having low dietary diversity.

### Covariates

We additionally measured sex (“male” or “female”); age (calculated as number of years from the date of birth at time of interview); education attainment (“none/some primary”, “primary”, “ secondary” and “post-secondary”); household size (total number of individuals living in the household at the time of the interview); work status (engaged in non-farm activities outside the home for income during the past 30 days); and wealth status (measured with an index computed with data on ownership of 20 household items and agricultural assets). We grouped participants into quartiles in accordance with the 2022 Kenya Demographic Health Survey ^(24)^.

### Statistical Analysis

We fit modified Poisson models to estimate the prevalence ratios (PR) for associated 95% confidence intervals (95%CI) for the association between participation in the agricultural intervention and dietary diversity. In the adjusted model, we controlled for age, marital status, education, work status, sex, household size, and wealth index, that were identified as minimally sufficient adjustment set using a directed acyclic graph (DAG). We evaluated sex as effect measure modifier by including an interaction term between sex and dietary diversity.

We addressed confounding using both covariate adjustment and Inverse Probability of Treatment Weights (IPTW), separately. Because these two approaches address confounding through different model assumptions, we wanted to gain a more robust understanding and handling of confounding by triangulating findings across the approaches. While we used an adjusted modified Poisson regression model to control for confounders directly in the model, the IPTW approach was used to predict treatment probabilities and to create a weighted sample in which the exposed and unexposed groups were more comparable. We generated IPTWs using the same set of covariates included in the adjusted Poisson model and applied these weights in both the overall analysis and in sex-stratified models. All analyses were carried out in RStudio using a significance level of 5%.

### Sensitivity Analyses

To assess for the potential influence of unmeasured confounding, we conducted a sensitivity analysis that calculated the E-value associated with the effect estimate for the agricultural intervention. The E-value estimates the minimum strength of association that an unmeasured confounder would need to have with both the exposure (agricultural intervention) and the outcome (dietary diversity) to fully explain away the observed association ^(25)^ . To contextualize the E-value magnitude, we compare it to the strength of the observed associations with measured covariates.

One concern we had was that exposure to the agricultural intervention could be tightly correlated with exposure to longer (and presumably more beneficial) durations of microfinance exposure. To determine whether the duration of microfinance membership was associated with dietary diversity, and isolate the effect of microfinance membership duration as exposure from that of an agricultural intervention, we conducted a sensitivity analysis by restricting the study sample to an agricultural intervention exposed and unexposed groups separately. We then used the duration of microfinance membership as exposure to identify whether it predicted dietary diversity. We compared the magnitude and direction of the point estimate with that for the overall association between an agricultural intervention and dietary diversity.

## RESULTS

### Baseline Characteristics

Of the 312 study participants in the overall sample, most participants (87%) were women, 52% were aged 30 years and above with a mean age of 40 years, 78% had not worked outside their homesteads for income during the last 30 days, and 85% were currently married. In terms of education attainment, most participants (46%) had primary level of education, compared to those in secondary (21%) and post-secondary education (10%). The median household size was 5 (IQR= 4, 7). There were some significant differences in those who did and did not receive the agricultural intervention (Table 1). Participants in the agricultural intervention were more likely to be in older age categories, be currently married, have post-secondary education attainment, be in the highest wealth quantile, and have worked outside home for income, compared to those not in the agricultural intervention. There were no significant differences in sex, work status, and household size between the two groups.

**Table 1:** Study population characteristics in the full sample and by agricultural exposure status (n=312)

|  |  |  |  |  |
| --- | --- | --- | --- | --- |
| **Agricultural Exposure Status** |  |  |  |  |
|  |  | **YES** | **NO** | **p value** |
| **Sociodemographic characteristics** | **Overall sample (100%)** | **N = 156** | **N=156** |  |
| **Sex** |  |  |  | 0.24 |
| Male | 40 (13%) | 24 (15%) | 16 (10%) |  |
| Female | 272 (87%) | 132 (85%) | 140 (90%) |  |
| **Age Category** |  |  |  |  |
| <30 | 32 (10%) | 6 (4%) | 26 (17%) | <0.0001 |
| 30-39 | 106 (34%) | 47 (30%) | 59 (31%) |  |
| 40-59 | 125 (40%) | 69 (44%) | 56 (36%) |  |
| 60 + | 49 (12%) | 34 (22%) | 15 (9%) |  |
| **Marital status** |  |  |  | 0.02 |
| Never married | 46 (15%) | 33(21%) | 13(8%) |  |
| Currently married | 264 (85%) | 122 (79%) | 142(92%) |  |
| Missing | 2 | 1 | 1 |  |
| **Education** |  |  |  | 0.005 |
| None/Some Primary | 67 (21%) | 21 (13%) | 46 (29%) |  |
| Primary | 146 (46%) | 80 (51%) | 66 (42%) |  |
| Secondary | 67 (21%) | 35 (22%) | 32 (20%) |  |
| Post-secondary | 32 (10%) | 20 (12%) | 12 (8%) |  |
| **Worked outside home (last 30 days)** |  |  |  | 0.2 |
| Yes | 69 (22%) | 40 (26%) | 29 (19%) |  |
| No | 243 (78%) | 116 (74%) | 127 (81%) |  |
| **Household size [Median (IQR)]** | 5 (4,7) | 5 (4,7) | 5 (4,7) | 0.3 |
| **Wealth quartile** |  |  |  | 0.006 |
| Q1(lowest) | 84 (27%) | 35 (22%) | 49 (31%) |  |
| Q2 | 70 (22%) | 27 (17%) | 43 (28%) |  |
| Q3 | 81 (26%) | 49 (31%) | 32 (21%) |  |
| Q4 (Highest) | 70 (22%) | 42 (27%) | 28 (18%) |  |
| Missing | 7 | 3 | 4 |  |
| **Household Dietary Diversity** |  |  |  | <0.0001 |
| Yes | 168 (54%) | 104 (67%) | 64 (41%) |  |
| No | 143 (46%) | 52 (33%) | 91 (59%) |  |

### Agricultural Intervention and Dietary Diversity

Participants who received the agricultural intervention self-reported higher dietary diversity (67%) compared to those not in the agricultural intervention (46%). This was reflected in the unadjusted regression results [PR (95% CI): 1.61 (1.31, 2.03), Table 2]. Adjustment for wealth status, household size, education attainment, and sex slightly attenuated results towards the null but did not substantially change the magnitude of the point estimate [aPR (95% CI): 1.46 (1.04, 2.06)]. Results from the IPT weighted model were similar in magnitude and direction to the adjusted model [PR (95% CI): 1.42 (1.14, 1.78)]. There were no meaningful differences in effect sizes for men and women. In men, those with the agricultural intervention exposure had 32% higher rates of high dietary diversity, with confidence intervals overlapping the null (aPR (95% CI): 1.32 (0.35, 4.97)). In women, those with the agricultural intervention exposure had 49% higher rates of high dietary diversity, with confidence intervals excluding the null (aPR (95% CI): 1.49 (1.05, 2.13)). We found no effect measure modification by sex in our study sample (Wald p-value for interaction term=0.83).

**Table 2:**
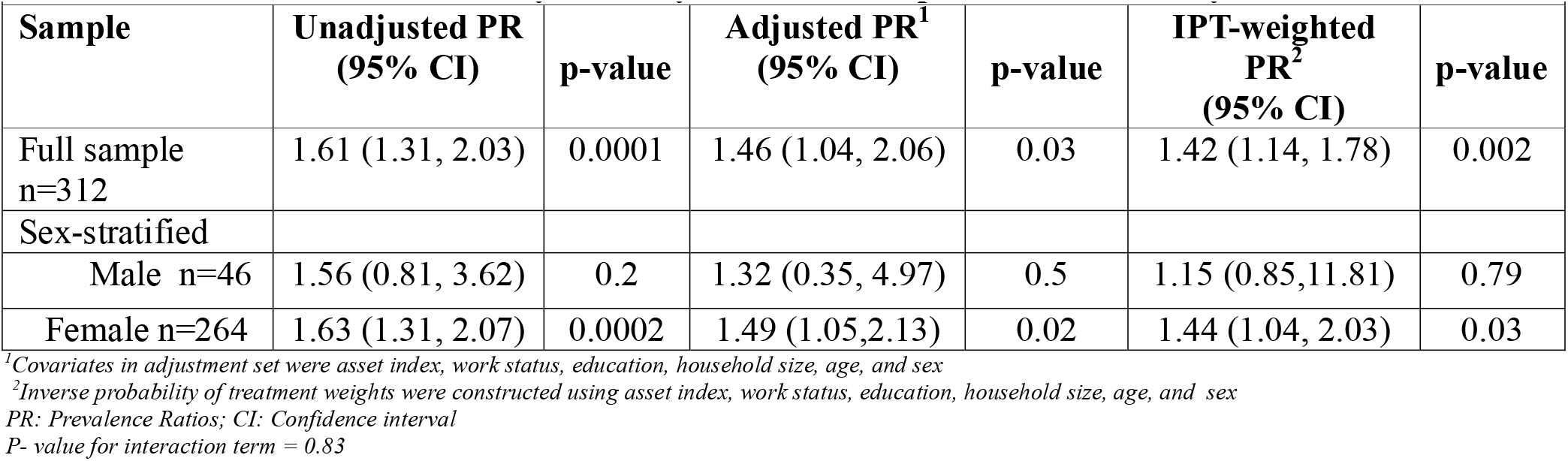
Prevalence ratios for the associations between exposure to an agricultural intervention and household dietary diversity in the full sample, and stratified by sex.

We calculated an E-value of 2.27 (95% CI lower bound: 1.42) corresponding to the magnitude of an effect an unmeasured confounder would need to have to fully attenuate the observed association (Table 3). Results from the sensitivity analysis showed that the magnitude of the point estimates for work status (aPR=0.95), age (aPR=1.0), wealth status (aPR=1.44), education (aPR=1.27), and household size (aPR=0.99) were smaller than 2.27, suggesting that the observed association is large enough that it is unlikely to be negated by unmeasured confounders.

Our analysis to isolate the effect of microfinance program exposure on dietary diversity from an agricultural intervention exposure supported our hypothesis that our results were largely driven by the agricultural intervention. We stratified our study sample into groups exposed and unexposed to the agricultural intervention, and used the duration of microfinance membership as the independent variable. Our results for the association between microfinance exposure and dietary diversity were smaller in magnitude than our primary findings, similar in magnitude between groups exposed and unexposed to the agricultural intervention, and not statistically significant in the exposed group (aPR (95% CI): 1.22 (0.28, 5.04) or the unexposed group (aPR (95% CI): 1.16 (0.75, 1.80) (Table 4)). By contrast, evidence that microfinance program maybe strongly associated with improved dietary diversity would have been demonstrated by larger, and statistically significant effect estimates for the association between microfinance program duration of exposure, and dietary diversity among participants unexposed to the agricultural intervention, suggesting that changes in dietary diversity was not driven by the timing of microfinance exposure.

## DISCUSSION

We found that exposure to the agricultural intervention was associated with higher levels of optimal dietary diversity among participants enrolled in group-based microfinance in our study area of rural Western Kenya. Our results suggest that integrating agricultural interventions into a group-based microfinance programs may address undernutrition among socioeconomically disadvantaged individuals by promoting access to diverse food groups in Kenya.

We found no evidence that the observed associations differed between men and women, suggesting gender-equitable nutritional impacts of the agricultural intervention although point estimates were smaller, and imprecise among men in our study sample. However, we need to acknowledge that our inability to detect effect measure modification by sex could also be explained by the low number of men in our study sample, and that our findings were not incompatible with a smaller effect size for men. The sex distribution we observed which skews strongly female reflects the real-world: women play a larger role in group-based microfinance programs with socioeconomic enhancement activities like agricultural interventions. Because findings from prior studies across similar settings indicate that women often assume primary responsibilities for food purchasing, preparation, and intra-household food allocation, along with additional burdens shaped by prevailing gender norms ^(26,27)^, it is possible that the nutritional impact of the agricultural intervention in our study settings would differ by sex. Future work should prioritize using larger samples to improve the precision of sex-specific estimates and to more rigorously evaluate potential differences by sex as no prior research has examined whether agricultural interventions affect men and women differently in these settings.

Our findings are consistent with prior research demonstrating improvements in optimal dietary diversity following an agricultural intervention. Although previous studies with consistent findings have not specifically examined agricultural interventions delivered through a group-based microfinance programs, many evaluations of agricultural interventions programs designed to improve nutrition in low- and middle-income countries have generally reported increases in household dietary diversity among participating households ^(9–14,16–18,28)^ .

However, our findings contrast with prior work that found no association between agricultural interventions and dietary diversity in Vietnam and Zambia ^(17,18)^. One possible explanation for the contrast is that the study in Vietnam was conducted in a setting where baseline dietary diversity was already high, potentially limiting the intervention’s ability to further increase dietary diversity, while an agricultural intervention in Zambia promoted farm production diversity of three food groups only: legumes, pulses, and nuts intended for sale and household consumption. Consequently, there were few participants who reported consumption of more than three food groups in the intervention arm, compared to the control group, unlike in our study where the majority of participants exposed to the agricultural intervention reported consumption of all five food groups considered adequate for optimal dietary diversity standards.

There are several plausible pathways through which an agricultural intervention may improve dietary diversity. First, an agricultural intervention may promote dietary diversity through agricultural production by households which increases the availability of different food groups for household consumption. In particular, research on agricultural interventions that focus on planting of a variety of indigenous vegetables and livestock rearing reported increased consumption of nutrient-dense foods in Nepal, Rwanda, and Myanmar ^(14–16)^. Second, an agricultural intervention may promote dietary diversity through the sale of agricultural produce which generates additional household income that can be used to purchase more food groups, thereby improving diverse diets in the households. In particular, current evidence from multiple settings indicate that crop and livestock production, and income from sale of agricultural produce empowers participating household members to purchase a variety of food groups, resulting in improved dietary diversity ^(29– 31)^. Third, an agricultural intervention may strengthen women’s empowerment by increasing their control over agricultural production. This mechanism has been demonstrated by prior research that found that empowered women with control, and greater decision-making power regarding food purchases resulted in improved household dietary diversity ^[32,33]^ .

Given the cross-sectional nature of this study, our findings should be interpreted with caution. First, the timing of data collection period for the current study coincided with the harvest season when foods were plentiful and accessible. As a result, dietary diversity may have been artificially elevated across both groups. However, we minimized this bias by restricting our data collection timing to the same season for both exposure groups to improve validity of the measure of association for that season. Future work may implement longitudinal studies during lean and harvest seasons to better capture typical dietary patterns and assess the agricultural intervention effect across additional seasons. Secondly, the cross-sectional study design may have limited our ability to establish temporal association between the agricultural intervention and dietary diversity. To partially address this concern, we framed survey questions to be in reference to specific time periods for the agricultural intervention exposure (6 months) and dietary diversity outcome (24 hours). However, the possibility that those who already had higher dietary diversity were more motivated and/or able to engage in the agricultural training remains and should be a focus for future work with randomized designs.

As participation in the agricultural intervention and its association with dietary diversity maybe strongly influenced by socioeconomic and demographic factors, confounding was a major concern in our study. Unlike most earlier studies examining the relationship between agricultural interventions and dietary diversity, our analysis addressed confounding by using DAG to identify the minimally sufficient set of covariates for adjustment, ensuring these variables were carefully measured, and controlled for in the modified Poisson model. We also applied IPTW using the same set of covariates to provide a complementary estimate, recognizing that IPT-weighted results might differ from modified Poisson regression estimates. However, the IPT-weighted findings derived from a weighted sample in which the distribution of measured confounders was balanced between the exposed and unexposed groups were similar in the magnitude and direction to the modified Poisson regression results, further strengthening the validity of our findings. We also calculated E-values to assess the impact of unmeasured confounding and conducted a sensitivity analysis restricting our study sample by the agricultural intervention’s exposed and unexposed groups separately in order to control for microfinance duration. Altogether, triangulating across results, we find support that our results are robust to potential bias from measured and unmeasured confounding. But, of course, the possibility that our results may be driven by uncontrolled confounding still exists and future work should consider more rigorous confounding control efforts using either randomized controlled trials or natural experiments.

The generalisability of our study findings to other populations who are not members of a group-based microfinance program is untested and therefore is limited. However, the pathways (socioeconomic pathways) through which agricultural interventions may be associated with improved dietary diversity may hold true across populations in similar settings supporting cautious generalisability our study results. Notwithstanding, our study contributes new evidence on the positive association between participating in the agricultural interventions delivered through a group-based microfinance program and dietary diversity from a Kenyan perspective.

## CONCLUSION

This study contributes to the small but growing body of evidence that participating in the agricultural interventions was associated with improved dietary diversity and is, to our knowledge, the first to examine the association within a group-based microfinance program in resource-scarce settings. We present the first evidence of this association in rural Western Kenya. Future work using large samples should build on these findings and focus on designing longitudinal studies to establish temporal ordering and causal relationships, evaluate potential differences by sex, as well as the feasibility and sustainability of integrating the agricultural interventions into microfinance platforms to improve dietary diversity and nutrition outcomes in resource-scarce settings.

## Data Availability

All data produced in the present study are available upon reasonable request to the authors

## Acknowledgements

We thank the <u>**B**</u>ridging <u>**I**</u>ncome <u>**G**</u>eneration for grou<u>**P I**</u>ntegrated <u>**C**</u>are microfinance members who volunteered to participate in this study

## Authorship

**CO**: conceived the research idea, protocol development, funding acquisition, data collection, analysis and writing of the original draft; **JK**: project administration, and data collection; **JA**: Writing - review and editing; **SP**: Writing - review and editing; **RC**: conceptualization, methodology, review and editing, **NM:** methodology, review and editing; **CL**: conceptualization, methodology, review, and editing; **MR**; supervision, conceptualization, methodology, formal analysis, writing, review, and editing.

---

**Table S1:** Prevalence ratios for covariates and e-value for the agricultural intervention exposure.

P- value for interaction term = 0.83
|  |  |  |  |
| --- | --- | --- | --- |
| **Exposure variable:** | **aPR (95% CI)** | **E-value** | **Lower 95% CI** |
| Agricultural intervention | 1.46 (1.04, 2.06) | 2.27 | 1.24 |
| **Covariate adjustment set** |  |  |  |
| Work status | 0.95 (0.72, 1.23) |  |  |
| Household wealth | 1.44 (1.02, 2.05) |  |  |
| Age | 1.0 ( 0.99, 1.01) |  |  |
| Household size | 0.99 (0.94, 1.05) |  |  |
| Education | 1.27 (0.76, 2.07) |  |  |

E-value: Expected values, 95% CI: confidence interval, aPR: adjusted prevalence ratio

**Table S2:**
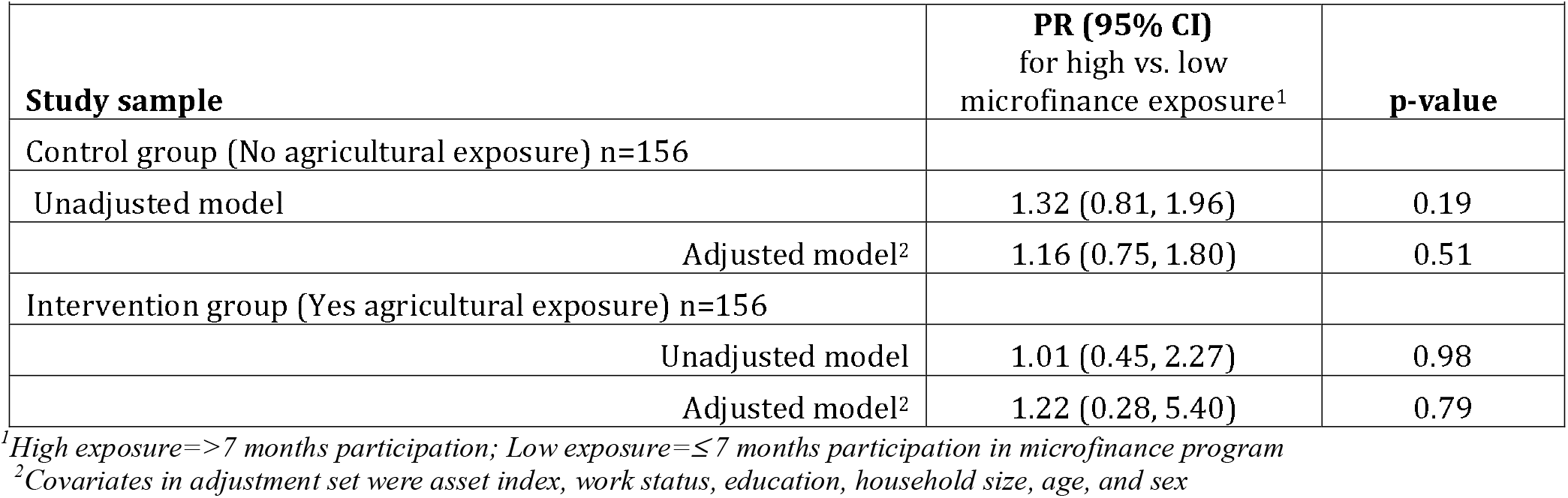
Prevalence ratios for the association between microfinance program and dietary diversity, stratified by exposure to agricultural intervention.

E-value: Expected values, 95% CI: confidence interval, aPR: adjusted prevalence ratio
|  |  |  |
| --- | --- | --- |
| **Study sample** | **PR (95% CI)**<br>**for high vs. low**<br>**microfinance exposure<sup>1</sup>** | **p-value** |
| Control group (No agricultural exposure) n=156 |  |  |
| Unadjusted model | 1.32 (0.81, 1.96) | 0.19 |
| Adjusted model<sup>2</sup> | 1.16 (0.75, 1.80) | 0.51 |
| Intervention group (Yes agricultural exposure) n=156 |  |  |
| Unadjusted model | 1.01 (0.45, 2.27) | 0.98 |
| Adjusted model<sup>2</sup> | 1.22 (0.28, 5.40) | 0.79 |

<sup>1</sup>High exposure=>7 months participation; Low exposure=<=7 months participation in microfinance program
<sup>2</sup>Covariates in adjustment set were asset index, work status, education, household size, age, and sex

